# Results of a nationally representative seroprevalence survey of chikungunya virus in Bangladesh

**DOI:** 10.1101/2024.03.25.24304711

**Authors:** Sam W. Allen, Gabriel Ribeiro Dos Santos, Kishor K Paul, Repon Paul, Ziaur Rahman, Mohammad Shafiul Alam, Mahmudur Rahman, Hasan Mohammad Al-Amin, Jessica Vanhomwegen, Taylor Smull, Kyu Han Lee, Emily S. Gurley, Henrik Salje

## Abstract

Chikungunya virus (CHIKV) is responsible for a rapidly increasing but poorly understood infection burden globally. Bangladesh experienced its first reported outbreak in 2008. Despite a number of subsequent isolated outbreaks, culminating in an enormous nationwide epidemic in 2017, very little is known about the burden or dynamics of chikungunya within the country, and the risk factors for infection. We conducted a nationally representative seroprevalence survey in 2016 in 70 randomly selected communities across the country. Individuals provided blood samples, which were tested for the presence of IgG antibodies to CHIKV. We also trapped and speciated mosquitoes. We found that 69/2,938 (2.4%) of individuals were seropositive to CHIKV. Seropositive individuals were concentrated in the centre and south of the country. We found that being seropositive to dengue virus (aOR 3.11 [95% CIs: 1.17 – 24.45]) and male sex (aOR 0.29 [95% CIs: 0.01 – 0.96]), were significantly associated with CHIKV seropositivity, however, *Aedes* presence, income, and travel history were not. Using a spatial prediction model, we estimate that at the time of the study, 4.99 million people in the country had been infected with CHIKV. These findings highlight high population susceptibility prior to the major outbreak in 2017 and that historic outbreaks must have been spatially isolated.

## Introduction

Chikungunya virus (CHIKV) is an *Aedes-transmitted* arbovirus first detected in Tanzania in 1953 which has since spread to the rest of Africa, Asia and South America, as well as sporadic outbreaks elsewhere [1]. The hallmark symptoms of chikungunya are abrupt onset of fever and joint pain [2,3], commonly accompanied by a rash [4]. The clinical presentation of chikungunya in humans is similar to that of dengue and a range of other illnesses, so is often misdiagnosed [2,5–9]. Around half of infected individuals suffer long-term effects, the most common of which is arthralgia [10–15] that can continue for months or even years post-infection, severely decreasing quality of life [16–18]. High attack rates coupled with frequent severe, persistent symptoms means that CHIKV outbreaks can place a large burden on communities, especially in lower-middle-income countries [2].

Limited access to testing and misdiagnoses mean that entire CHIKV epidemics are frequently missed, especially in lower-middle-income countries [19,20] and we rarely have a good understanding of the underlying burden from CHIKV in any affected setting, as well as individual-, household- and population-level predictors of infection risk. This critical knowledge gap means we do not know where to appropriately target interventions. This is becoming increasingly relevant, with the licensure of the first chikungunya vaccines expected in the near future and the demonstration that the targeted release of *Wolbachia*-infected mosquitoes can reduce incidence [21,22]. In this context, seroprevalence studies can help, especially as CHIKV infection appears to result in long-lasting and immunising antibodies[23]. By measuring the presence of antibodies in the population, we can quantify the underlying level of infection history in a population[20]. Further, by combining the results of seroprevalence studies from multiple locations with mathematical models, we can estimate the burden across the population, and the changing level of immunity [24].

Here we focus on Bangladesh. CHIKV was first identified in Bangladesh in 2008 during an outbreak in the northwest of the country in two villages near the Indian border [25]. Subsequently, there were localised outbreaks detected between 2011 and 2016 [2,26]. In 2017, Bangladesh experienced a far larger outbreak, with infections reported nationwide with as many as a million reported cases in total [2,27,28]. However, it remains unclear if there was substantial transmission across the country prior to the large outbreak in 2017. In this project, we present the results of a nationally representative seroprevalence study from Bangladesh. We visited communities around the country in 2016, allowing us to quantify the level of transmission, identify risk factors for infection, and the level of immunity prior to the major outbreak.

## Results

We visited 70 randomly selected communities in Bangladesh. We collected blood and administered questionnaires from 2,938 individuals. Simultaneously, we collected mosquitoes using BG-Sentinel traps during our survey. There was a mean of 42 participating individuals per community (range 39-57), and a mean of 10 participating households per community (range 10-12). The mean age of participants was 30 and 52% of participants were female. Our IgG Luminex-based antibody assay divided individuals into seropositive and seronegative individuals (Figure S1). Among the participants, 2.4% were seropositive to CHIKV, with all seropositive individuals coming from 16 communities (23% of all communities), concentrated in the central and south of the country. Among seropositive communities, mean seropositivity was 10.45%, ranging from 2.08 to 39.02%. Notably, a single individual from the 3 communities in Dhaka city were seropositive. Seropositivity was largely consistent across different age groups, except for those aged under five years in age, who had a seropositivity of 0% compared to 2.4% for those over five years (p-value 0.26) (Table 1). We used logistic regression to identify covariates associated with being seropositive. We compared models with or without a spatial covariance term, as well as models with or without household and community level random intercepts. The best fitting model, as measured by WAIC (Watanabe-Akaike information criterion, a measure for model comparison), included household and community level random intercepts, however, covariate estimates were largely consistent across the different models considered (Figure S2).

**Table 1:**
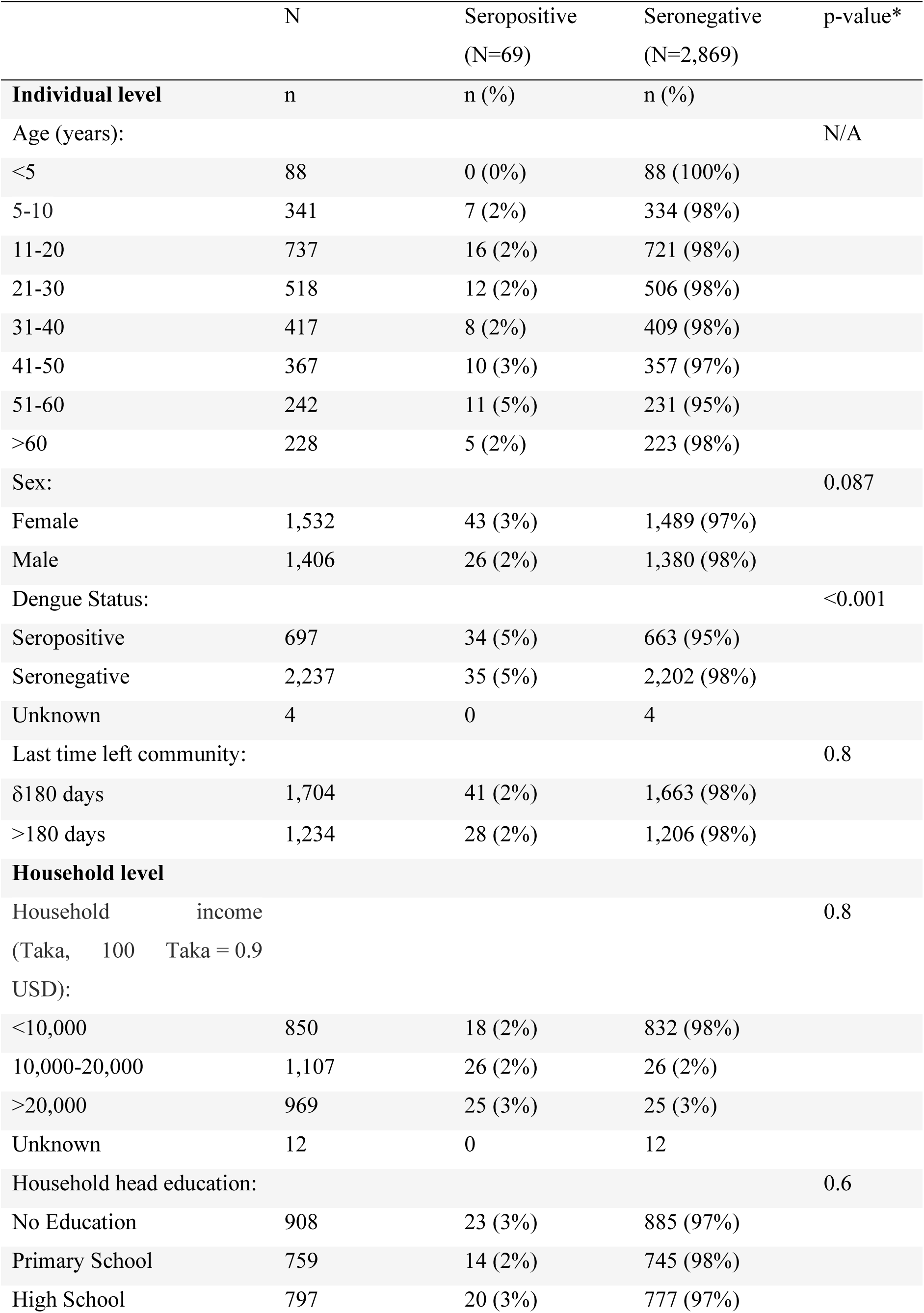
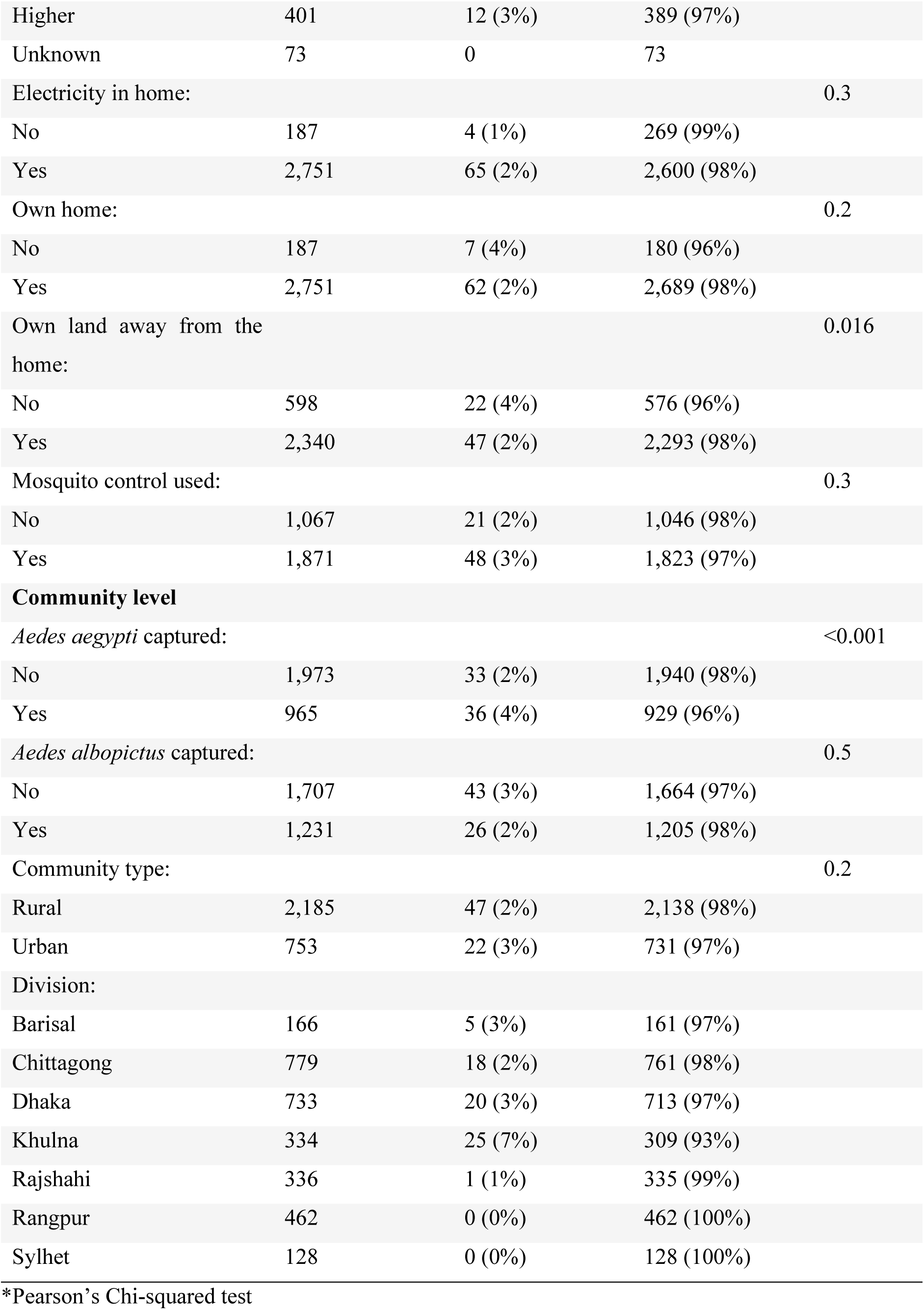
Individual-, household- and community-level characteristics of participants across Bangladesh, stratified by serostatus to chikungunya in 2015/16. Seropositivity determined based on the presence of IgG antibodies against CHIKV.

**Table 2:**
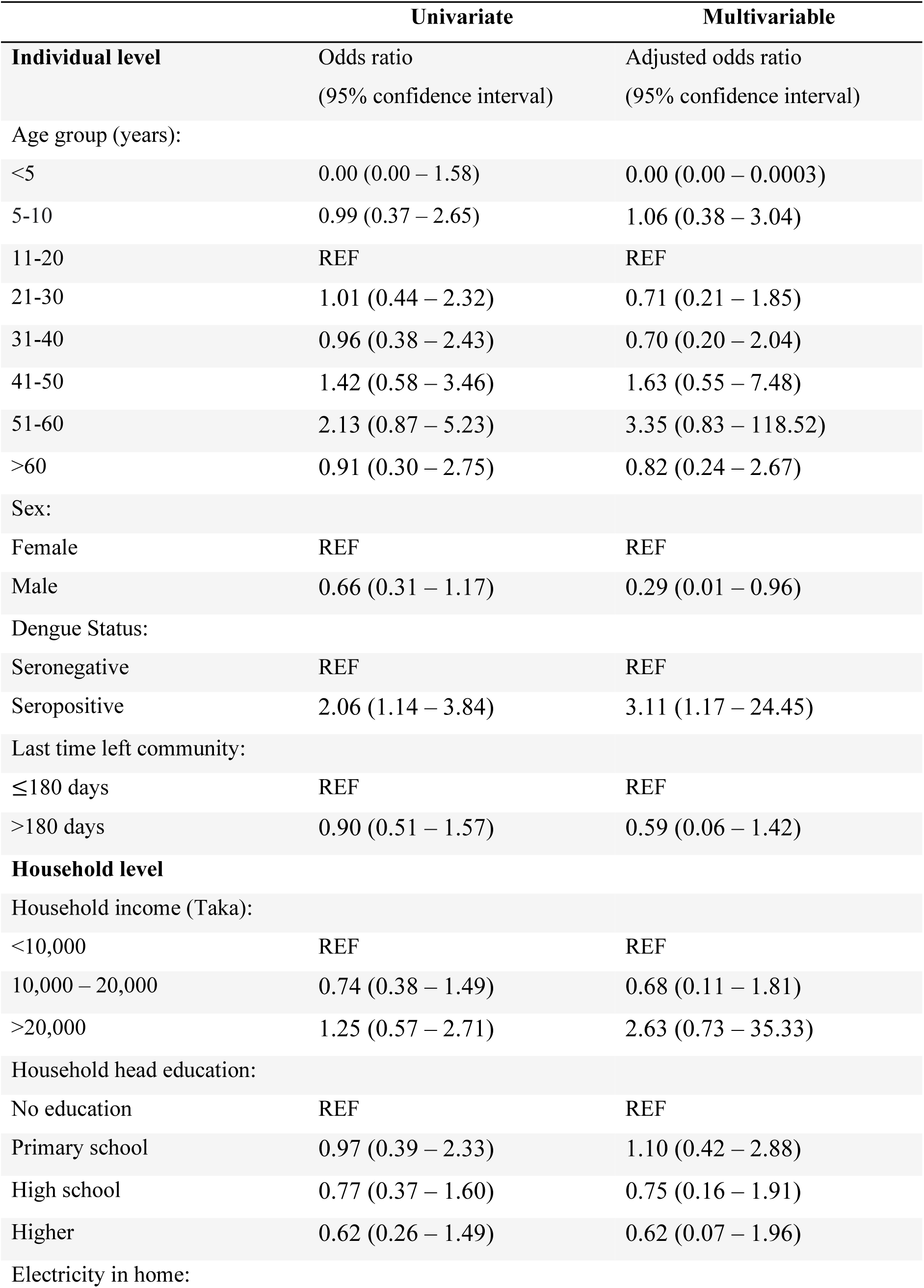
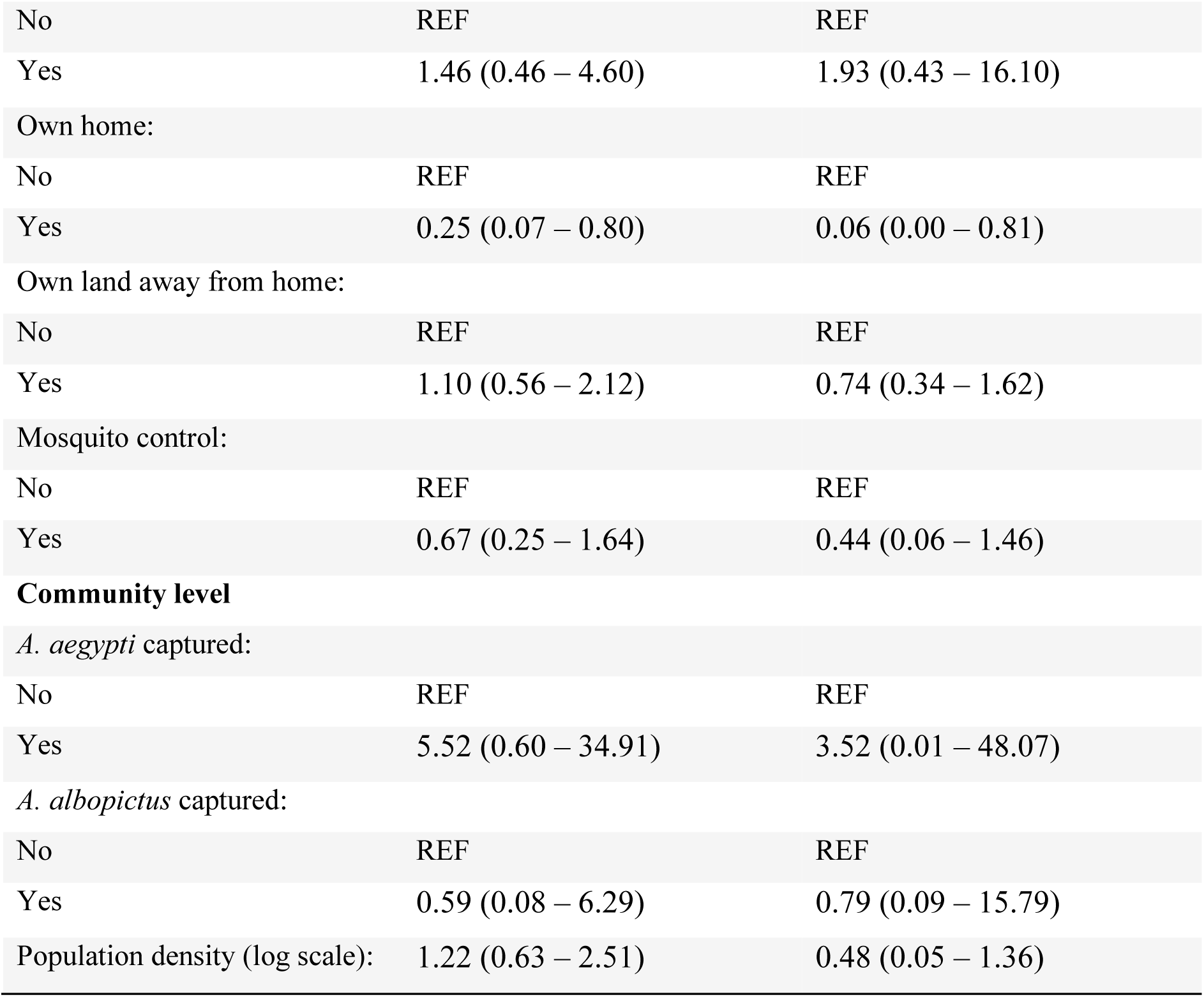
Results of univariate and multivariable logistic regression. The multivariable model selected included random community and household intercepts, but no spatial field (Model 5), on the basis of WAIC (Table S1).

We compared a suite of models to identify individual-, household- and community-level covariates linked to CHIKV seropositivity. These models either included or did not include household and community random effects and a spatial field. The best fitting model had both household and community random effects but no spatial field (Table S1). In this model, we found most factors were not associated with CHIKV seropositivity. This includes the presence of *Aedes aegypti* and *Ae. albopictus* in the community, population density, household income and travel history. However, being aged under 5 years (aOR: 0.00, 95% CI: 0.00 – 0.0003), being seropositive for dengue virus (aOR: 3.11, 95% CI: 1.17 – 24.45), male sex (aOR: 0.29, 95% CI: 0.01 – 0.96) and household ownership (aOR: 0.06, 95% CI: 0.00 – 0.81) were significantly associated. Either *Ae. aegypti* or *Ae. albopictus* was found in 46 communities (66% of communities) (Figure S3).

We next explored whether within the communities where CHIKV seropositivity was detected, living with a seropositive individual was a risk factor for being seropositive. We found that within these communities, individuals who lived with a seropositive householder member had 2.80 (95% CIs: 1.47 – 4.85) times the probability of being seropositive as compared to individuals living with only seronegative individuals.

To estimate the overall seropositivity across the country, we used a spatial prediction model. We used the estimated population distribution in the country to identify the number of infected individuals. Overall, we estimate that 4.99 million people (95% CI: 4.89 - 5.08 million) in Bangladesh had been infected with chikungunya at some point in their lives as of 2016, with the highest risk concentrated to a few focal hotspots (Figure 2A). This equates to about 2.49% (95% CI: 2.45 – 2.54%) of the national population, consistent with the estimate produced using the crude proportion seropositive among individuals in the serosurvey (2.35% of the population, or about 4.70 million individuals). To validate our spatial prediction model, we removed all data from a subset of individual communities in turn from our model and used the remaining data to fit a new model. We then used the fitted model to predict in the removed locations. We found we could accurately predict seropositivity in the removed locations (Pearson correlation of 0.95) (Figure 2B).

**Figure 1:**
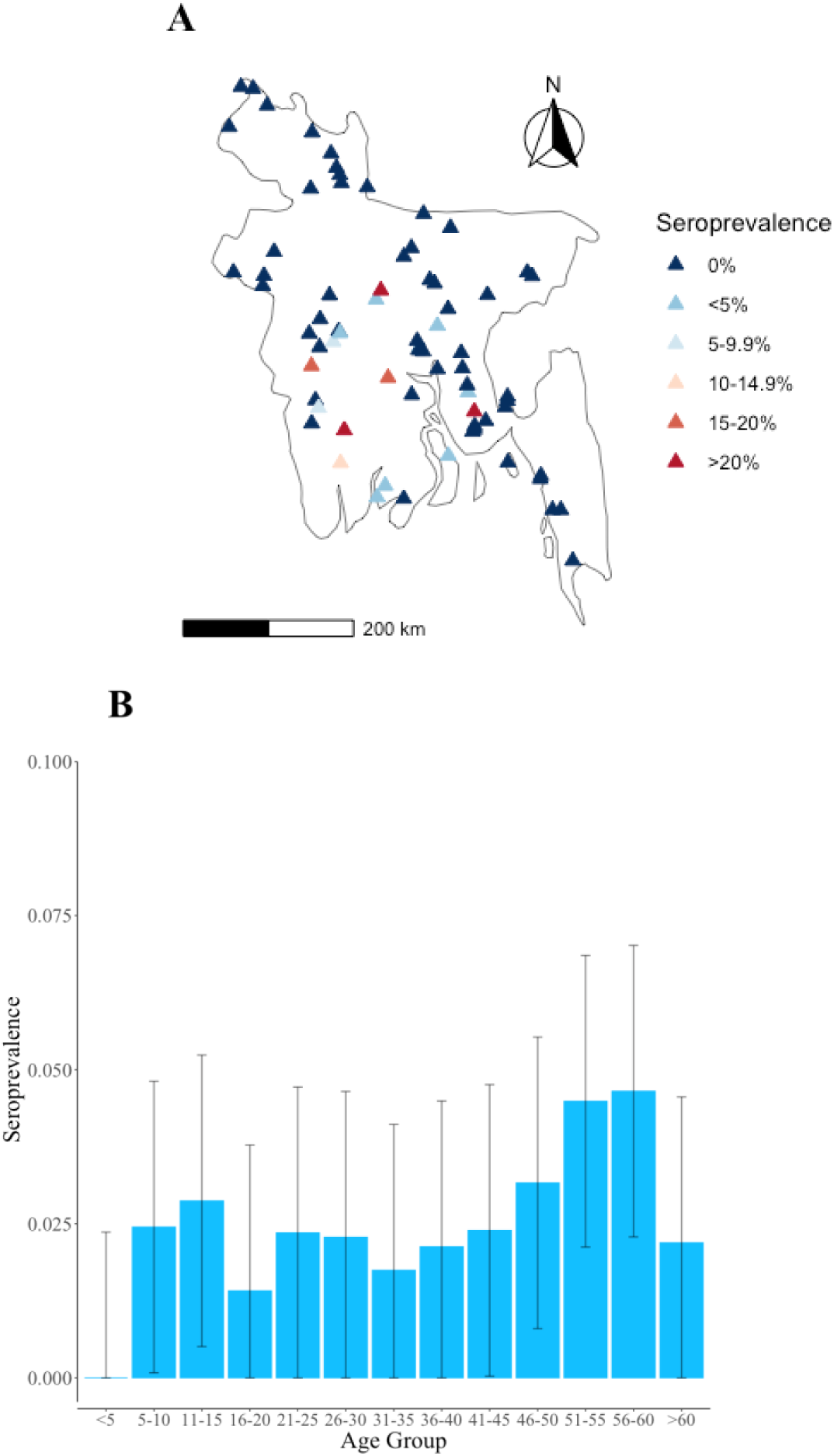
Proportion seropositive. **(A)** Map of sampled communities and proportion seropositive to CHIKV. **(B)** Proportion seropositive by age.

**Figure 2:**
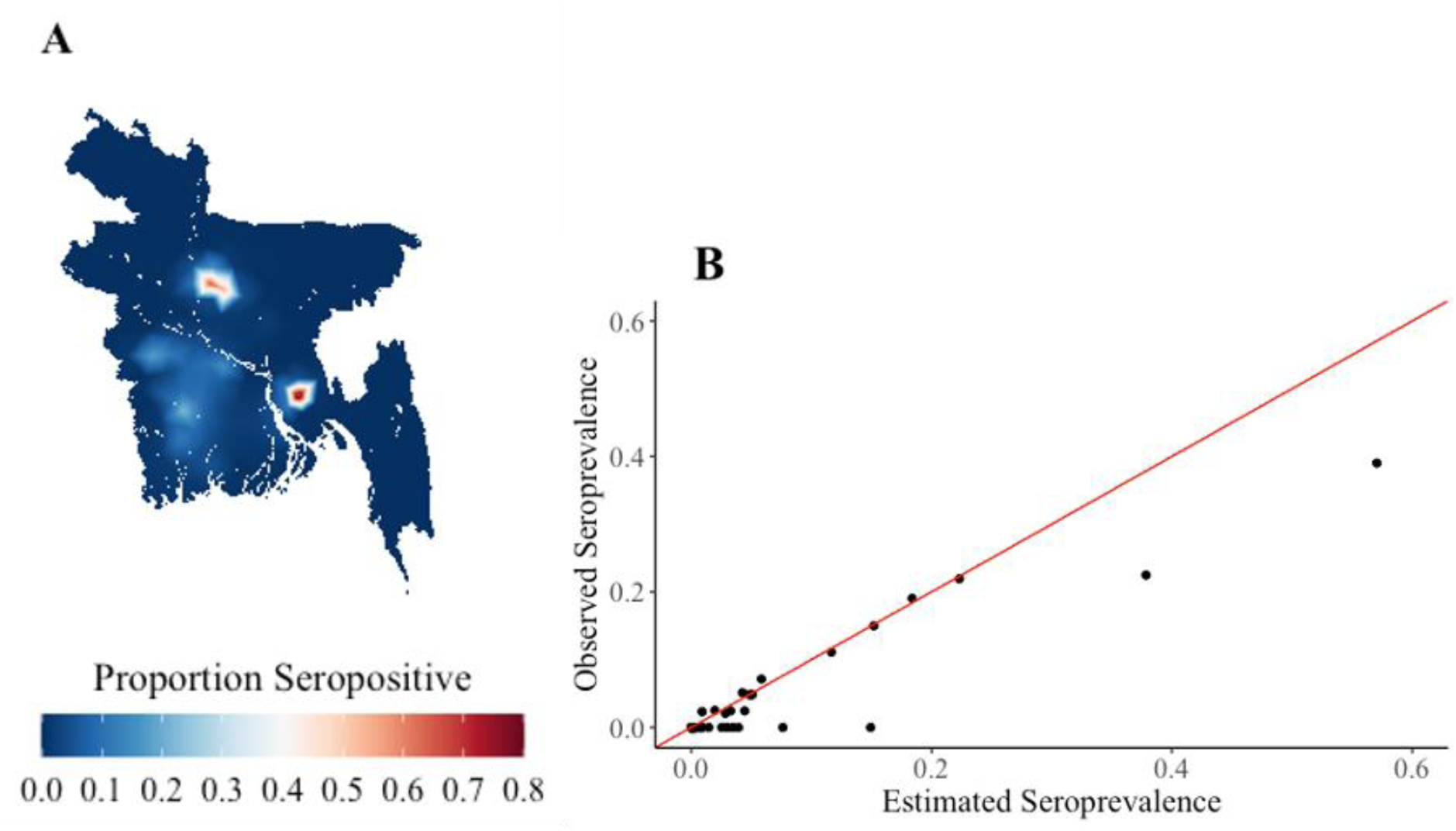
Estimated map of seropositivity. **(A)** Modelled seropositivity in Bangladesh in 2016 **(B)** Held out cross validation where communities were removed from model fitting process and the rest of the data used to fit models. The plot shows the comparison with the observed versus the predicted in the removed locations.

## Discussion

The results from this first nationally-representative serosurvey of CHIKV infection in Bangladesh demonstrate that by 2016, CHIKV had been present in parts of the country, especially the South. Overall, only a relatively small proportion of the population, representing around 5 million individuals, had previously been infected. The high level of population susceptibility at this time can help explain the magnitude and spatial extent of the subsequent major outbreak in 2017.

CHIKV seropositivity was relatively constant across age groups, which indicates that individuals of all ages have had the same cumulative exposure to risk of infection. This is indicative of a recent emergence of CHIKV in Bangladesh, though the decreased seropositivity among those aged under five suggests limited exposure in the years immediately preceding the serosurvey. We explored a wide range of individual-, household- and community- level risk factors to identify drivers of infection risk. Being seropositive to dengue virus, another virus transmitted by the same vectors, was an important predictor. This highlights the overlapping risk across Aedes-transmitted arboviruses, as previously identified elsewhere [29]. We identified a strong effect of the household, with individuals much more likely to be seropositive if they lived with other seropositive individuals. This finding is consistent with previous findings from Bangladesh and elsewhere that have identified the limited flight range of the vector as driving household infection risk, as biting typically occurs in the peridomestic environment [30]. A strong correlation of serostatus by household has also been observed with dengue virus [31]. Sex was another notable predictor, consistent with results from both Bangladesh and elsewhere that have consistently shown infection risk is higher among females [19,30,32–34]. It has been suggested that differences in mobility patterns may explain this increased risk, with women in Bangladesh spending more time in and around the home where mosquitoes reside [30].

Our results suggest that while prior to 2017 CHIKV outbreaks in Bangladesh have been spatially constrained, there was always the risk of a widespread epidemic. It remains unclear why prior outbreaks in the country died out without spreading widely. We identified either *Ae. aegypti* or *Ae. albopictus* mosquitoes in most communities, suggesting that conditions were suitable for transmission across the country. Introductions may have previously been in rural communities, which are less connected to urban hubs, and potentially died out from entering cooler parts of the year [30]. As population mobility continues to increase, we can expect even wider spread of both *Aedes* vectors and a concurrent increase in arbovirus outbreak risk [35].

This project highlights the utility of nationally representative seroprevalence studies, especially when combined with mathematical models. Using a sampling frame of all communities in Bangladesh allows us to generalise to the wider country. These same samples were used to create risk maps for a wide range of other pathogens, including cholera, dengue, and hepatitis E [24,36,37]. Further, the increased use of multiplex serology allows the parallel testing of multiple pathogens, maximising insights from individual blood draws, and limiting the need for numerous freeze-thaw cycles.

We note that our modelled estimate of seropositivity at the national level was very consistent with the crude level of seropositivity in our sample set (2.49% vs 2.35%). It is certainly possible that we did not sample communities affected by localised outbreaks but those outbreaks would not markedly change our estimates for the overall population level immunity for Bangladesh. Selection bias may have arisen in that individuals who were away from communities during visits, and hence more likely to travel frequently, may not have been able to participate. However, to minimise this risk, the study team arranged to visit households again when members were expected to return from travel. The travel covariate is also limited in that the questionnaire asked about most recent travel outside the community, which does not provide information on frequency, reason or destination of travel. All of these could be relevant to CHIKV infection risk.

In conclusion, we demonstrate high CHIKV susceptibility across Bangladesh prior to the major outbreak in 2017, and that prior outbreaks were largely spatially isolated in nature. Given the potential for large outbreaks, Bangladesh should be prioritised for new interventions, such as vaccines and *Wolbachia*-based vector control, as they become available.

## Methods

### Data collection

The protocol of the study has been described elsewhere [24]. Briefly, to obtain a nationally-representative sample, 70 of the 97,162 communities listed in the 2011 census were selected at random, with the likelihood of selection being proportional to population size. Each community was visited by the study team, who spent at least five days within each community. Visits occurred during October 2015 to January 2016. A further visit was made to communities where no *Aedes* had been previously trapped, during June and July 2016 for additional mosquito collection. Communities where Aedes were still not found after the second visit were defined as having an absence of *A. aegypti* and *A. albopictus*. The study team randomly selected at least ten households from each community. The heads of selected households were informed of the study and invited to participate. If they agreed, all other members of the household aged over six months were also invited to participate. Data collection was deemed to be complete for a community when at least 40 serum samples from at least 10 households were obtained.

The head of each participating household was led through a household questionnaire with a variety of questions regarding socio-economic status, such as education level, estimated household income and access to electricity. In addition, this questionnaire asked whether households had used any form of mosquito control in the last week and whether any member of the household owned land away from their home [33].

Each consenting household member (including the household head) was also guided through an individual-level questionnaire. If individuals were too young to answer this by themselves, an older household member was asked to answer on their behalf. These questionnaires covered demographic questions, such as age and sex, and also asked when participants had last travelled outside of the community [33].

All individuals who provided consent also had 5 ml of venous blood withdrawn by a phlebotomist. These blood samples were centrifuged and serum was then extracted separately and shipped in nitrogen dry shippers to icddr,b (International Centre for Diarrhoeal Disease Research, Bangladesh) laboratories in Dhaka. Individuals who were ill at the time of the survey were excluded from serum sampling. All serum samples were tested for antibodies against chikungunya to identify evidence of prior infection. This was done using a microsphere-based multiplex immuno-assay (MMIA) that measured the fluorescence intensity to both the recombinant E2 glycoprotein of the chikungunya virus and the background level of antibody activity at the individual level using a recombinant human O^6^-Methylguanine-DNA Methyltransferase protein (SNAP-tag). To determine CHIKV seropositive we first calculated the ratio between the fluorescence intensity to CHIKV and the control MGMT protein, and used a cut point of 5.5 to identify those with a history of CHIKV exposure (Figure S1). It has previously been estimated that a ratio of 5.5 on a linear scale (∼1.70 log scale) is the threshold for the MMIA to achieve 95% sensitivity and specificity (personal communication) [19].

### Regression analyses

We used the R-INLA package, which applies the INLA (Integrated nested Laplace approximation) method. INLA is a Bayesian approach to statistical inference for Gaussian Markov Random Field (GMRF) models [38]. A key benefit of INLA is that it can accommodate a range of GMRF models, including those with a spatial component. R-INLA allows these to be added to the model as random effects. This means that the spatial autocorrelation inherent in epidemiological data can be accounted for to isolate out the role of random spatial variation [39]. We modelled the dependence of two observations in this distribution using a covariance function, with the Matérn covariance function. R-INLA’s default priors were used beyond the setting of the spatial field, where a fixed smoothness parameter of 〈 = 2 was set. This represents a moderately smooth spatial field, and is a commonly selected value [40].

Covariates were divided into individual-level (age, sex, dengue serostatus and last time left community), household-level (income, highest education level achieved by head of household, electricity in home, own home, own land away from the home and use of mosquito control in the last week) and community level *(Ae. aegypti captured in the community, Ae. albopictus captured in the community,* division and log population density). Firstly, each covariate was included in a univariate logistic regression using R-INLA to assess individual relationships with serostatus. Random intercepts were also included for both the household and the community to account for correlation of observations within these sites. Following this, all covariates were included in a multivariable analysis. This generated an odds ratio and 95% confidence interval for each covariate from both univariate regression, and an adjusted odds ratio and 95% confidence interval from multivariable regression.

To explore the importance of the spatial correlation structure and the random household and community intercepts, additional models with different combinations of these included were also built. In total, five models were created:

1. The base model, featuring a Matérn spatial correlation structure, a random community intercept and a random household intercept.
2. A Matérn spatial correlation structure and a random household intercept only.
3. A Matérn spatial correlation structure and a random community intercept only.
4. A Matérn spatial correlation structure only.
5. Random household and community intercepts only.

Using the base model, the percentage of variance explained by each spatial correlation structure, random community intercept and random household intercept was determined. A model without any of these three was first run, and the variance calculated by taking the mean of the squared residuals. The three random effects were then added one-by-one. Each time, the variance was calculated and the percentage of the variance in the original model that the new model explained was estimated, to try and understand the impact of each addition. This was repeated both by adding the random community intercept before the random household intercept, and vice-versa.

### Household infection risk

To investigate whether living with a seropositive individual was a risk factor for being seropositive oneself, the risk ratio for living with a seropositive individual was calculated in the subset of communities with at least one seropositive individual. The risk ratio was then calculated as follows:

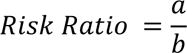

where,

a = the proportion of seropositive individuals living with seropositive individuals

b = the proportion of seropositive individuals living with seronegative individuals

This risk ratio was first calculated, and then bootstrapped for 1000 iterations to generate a distribution of estimates, from which a mean and 95% confidence intervals were extracted.

### Mapping chikungunya virus risk across Bangladesh

Seroprevalence by community was mapped by community to visualise the general spatial distribution of chikungunya in 2015/2016. Seroprevalence was defined as the number of individuals with detectable anti-chikungunya virus antibodies, expressed as a proportion of the total number of individuals surveyed in that community, calculated as follows:

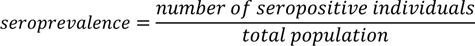

95% confidence intervals were also calculated for community seroprevalence using the Clopper-Pearson estimation method which is based on the exact binomial distribution (66). Seroprevalence was calculated for each community in the study and mapped to visualise spatial trends.

To explore infection risk across Bangladesh, a grid of 1 km x 1 km cells was placed over the country. A Bayesian framework featuring a Matérn spatial correlation structure was used to fit the model. Covariates from the multivariable regression could not be added to the spatial prediction because values for these covariates are not available for areas outside the study sites. Salje et al. [24] found that the inclusion of additional covariates (e.g. age and sex) beyond the spatial covariance term that can be obtained from demographic data did not markedly improve predictive accuracy, so these were not added to reduce unnecessary model complexity. The model was then fit to the 1 km x 1 km grid across the country to predict the seroprevalence in each of these cells.

To estimate the total number of people ever infected with CHIKV in Bangladesh at the time of the survey, the population density in each cell was multiplied by the fitted seroprevalence in each cell. Confidence intervals were generated by taking the 0.025 and 0.975 estimates from the model and applying the same technique.

To test the predictive performance model, a cross validation was performed. 1000 iterations were performed, with ten of the 70 communities left out during model fitting each run. The model was then used to predict the seroprevalence in the ten test communities. The mean predicted seroprevalence for each community was then used to generate an estimated seroprevalence, which was compared to the observed seroprevalence in each of the held out communities.

### Ethical clearance

The icddr,b and CDC ethical review boards approved of this study (protocol number PR-14058). All participating adults gave written informed consent. Children involved in the study had written, informed consent provided on their behalf by parents/guardians.

## Data Availability

All data produced in the present work are contained in the manuscript

## Footnotes

The authors would like to acknowledge funding from ERC 804744 and the CDC. The authors declare no conflicts of interest.

## Supplementary figures

**Figure S1.**
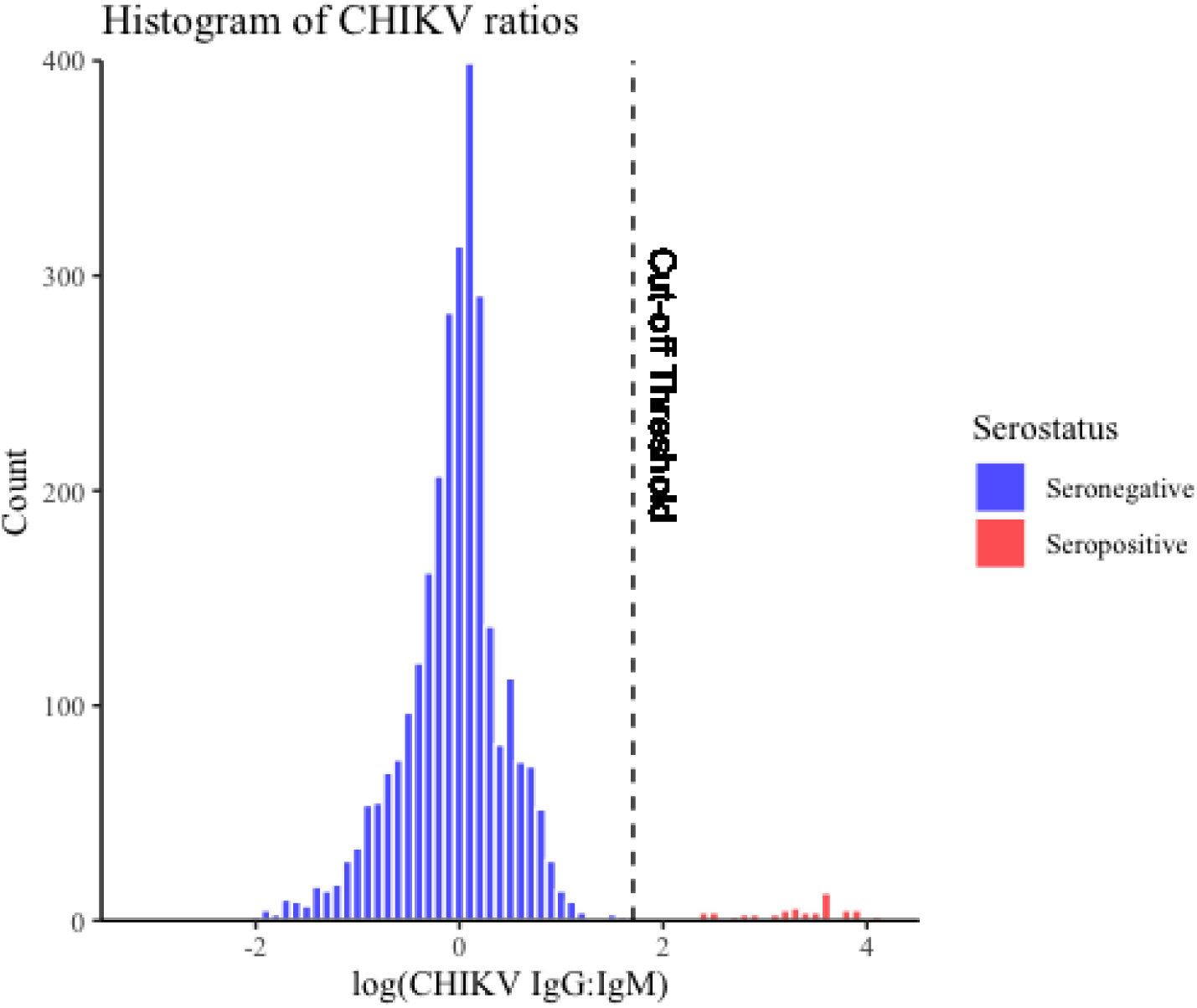
Histogram of the ratios between fluorescence intensity to CHIKV and the control SNAP-tag protein with the cutoff point of 5.5 marked (dashed line). Samples to the left of the dashed line are considered seronegative, and those to the right seropositive.

**Figure S2:**
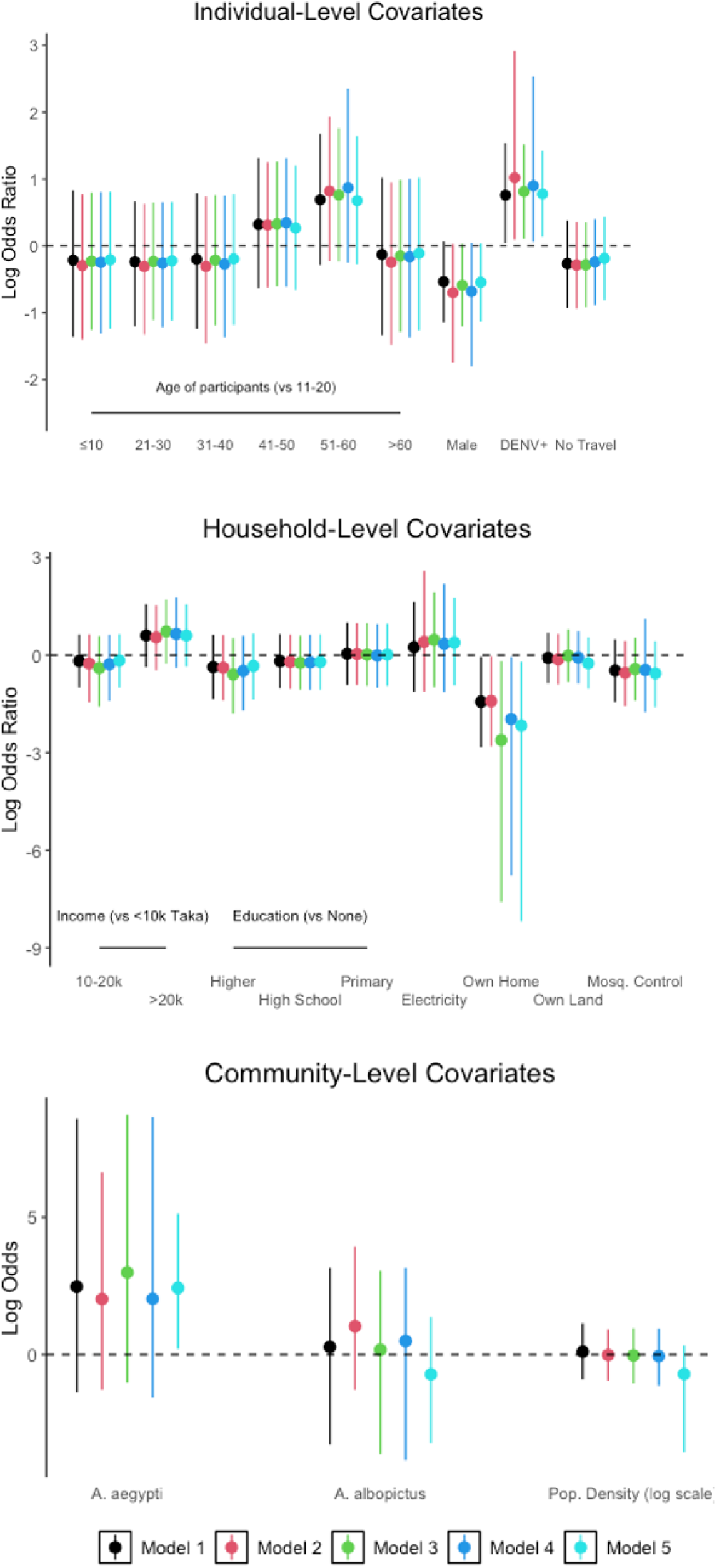
Difference in multivariable coefficient estimates. Run using logistic regression with a Matérn spatial correlation structure, random community intercept and a random household intercept (Model 1), a Matérn spatial correlation structure and random household intercept only (Model 2) a Matérn spatial correlation structure and random community intercept only (Model 3), a Matérn spatial correlation structure only (Model 4), and random household and community intercepts only (Model 5).

**Figure S3:**
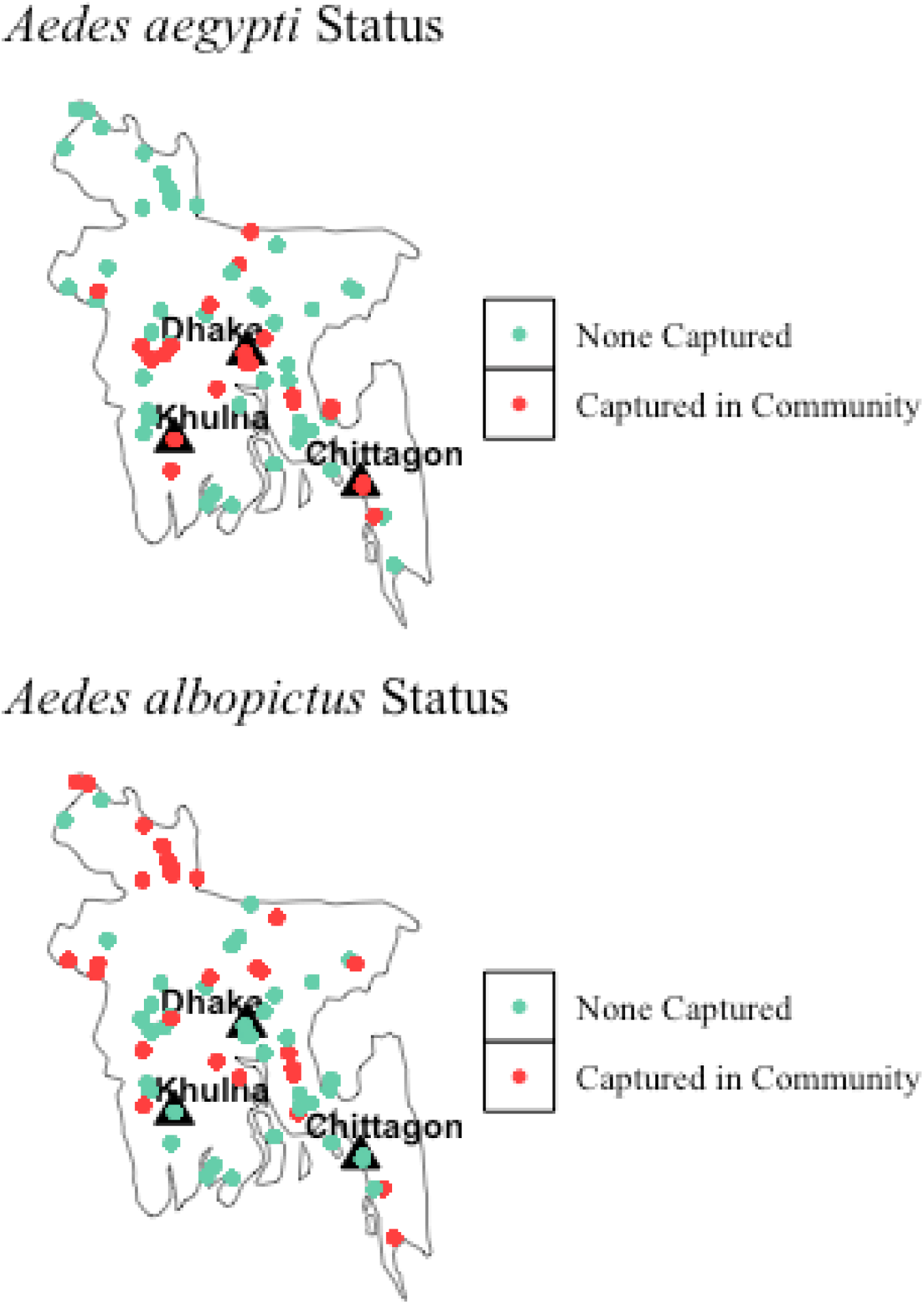
Presence of *A. aegypti* and *A. albopictus* in communities. The black triangles represent the three main cities in Bangladesh.

**Table S1:** Model comparison.

| Model | WAIC |
| --- | --- |
| 1 (Spatial correlation structure, community intercept and household intercept) | 2190.89 |
| 2 (Spatial correlation structure and random household intercept) | 3597.03 |
| 3 (Spatial correlation structure and random community intercept) | 2306.11 |
| 4 (Spatial correlation structure) | 3771.10 |
| 5 (Community and household random intercepts) | 496.22 |

**Table S2:**
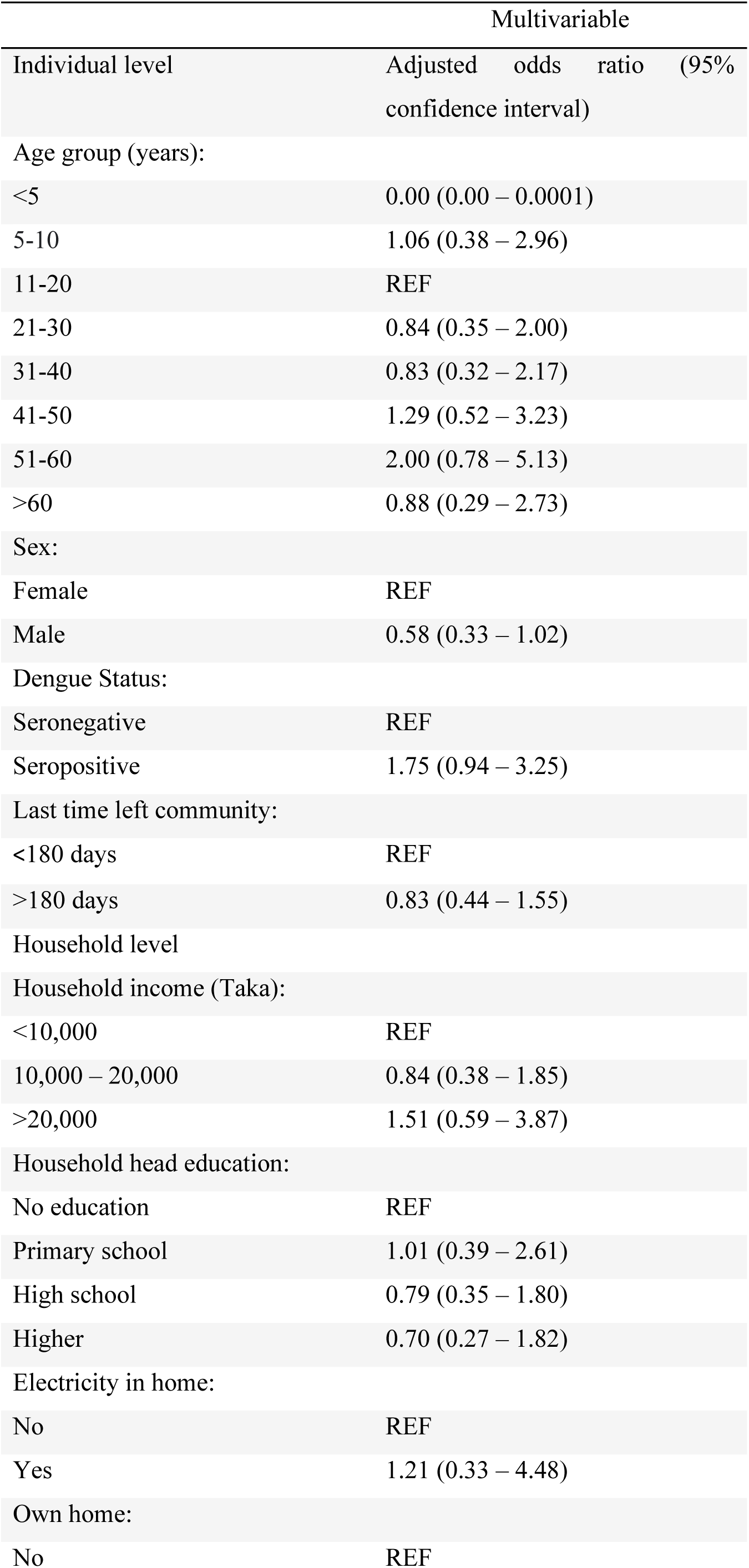
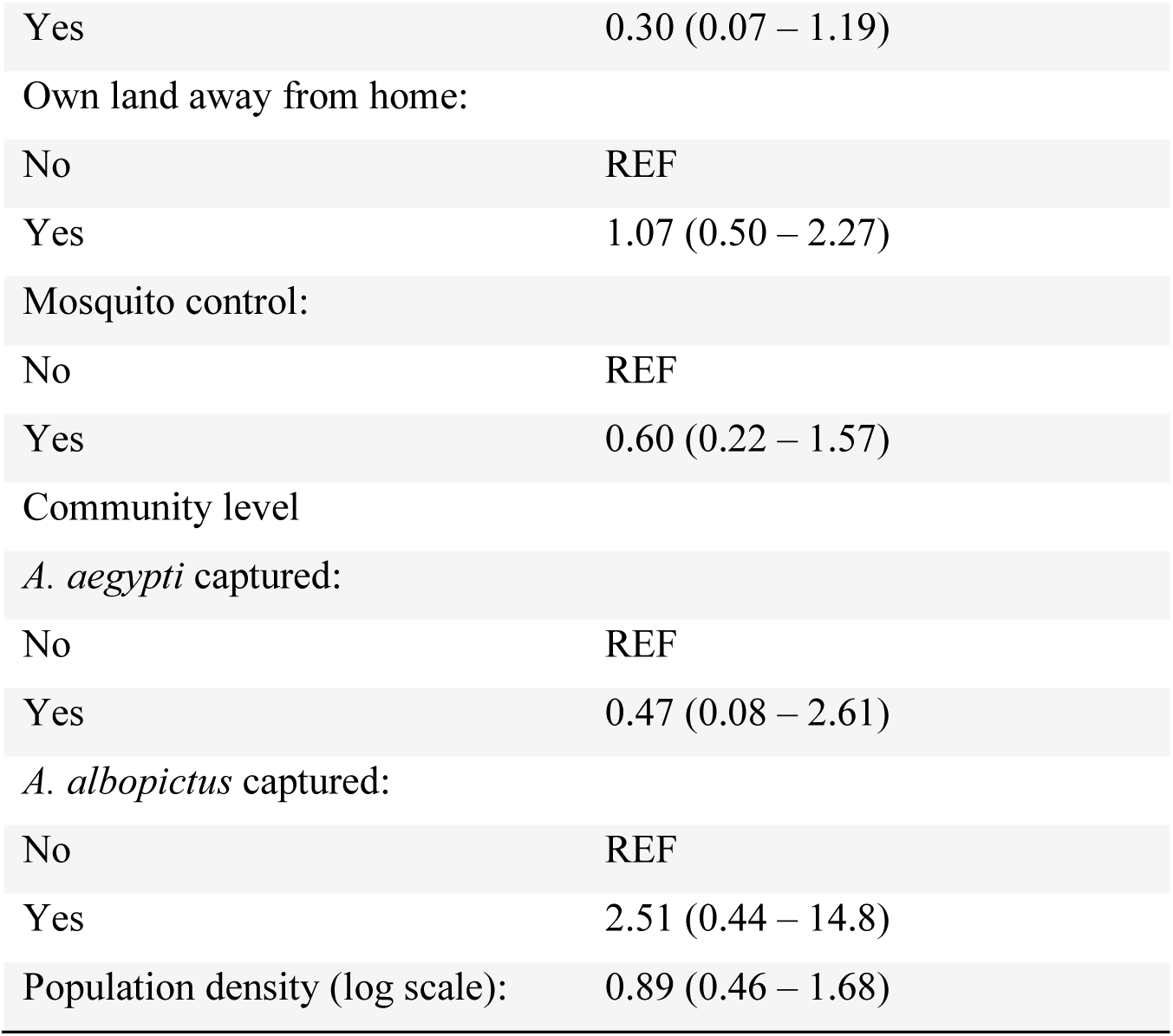
Results of logistic regression performed using data from the 16 communities with at least one seropositive individual.

| Multivariable |  |
| --- | --- |
| Individual level | Adjusted odds ratio (95% confidence interval) |
| Age group (years): |  |
| <5 | 0.00 (0.00 – 0.0001) |
| 5-10 | 1.06 (0.38 – 2.96) |
| 11-20 | REF |
| 21-30 | 0.84 (0.35 – 2.00) |
| 31-40 | 0.83 (0.32 – 2.17) |
| 41-50 | 1.29 (0.52 – 3.23) |
| 51-60 | 2.00 (0.78 – 5.13) |
| >60 | 0.88 (0.29 – 2.73) |
| Sex: |  |
| Female | REF |
| Male | 0.58 (0.33 – 1.02) |
| Dengue Status: |  |
| Seronegative | REF |
| Seropositive | 1.75 (0.94 – 3.25) |
| Last time left community: |  |
| <180 days | REF |
| >180 days | 0.83 (0.44 – 1.55) |
| Household level |  |
| Household income (Taka): |  |
| <10,000 | REF |
| 10,000 – 20,000 | 0.84 (0.38 – 1.85) |
| >20,000 | 1.51 (0.59 – 3.87) |
| Household head education: |  |
| No education | REF |
| Primary school | 1.01 (0.39 – 2.61) |
| High school | 0.79 (0.35 – 1.80) |
| Higher | 0.70 (0.27 – 1.82) |
| Electricity in home: |  |
| No | REF |
| Yes | 1.21 (0.33 – 4.48) |
| Own home: |  |
| No | REF |
| Yes | 0.30 (0.07 – 1.19) |
| Own land away from home: |  |
| No | REF |
| Yes | 1.07 (0.50 – 2.27) |
| Mosquito control: |  |
| No | REF |
| Yes | 0.60 (0.22 – 1.57) |
| Community level |  |
| *A. aegypti* captured: |  |
| No | REF |
| Yes | 0.47 (0.08 – 2.61) |
| *A. albopictus* captured: |  |
| No | REF |
| Yes | 2.51 (0.44 – 14.8) |
| Population density (log scale): | 0.89 (0.46 – 1.68) |

**Table S3:** Results of univariate and multivariable regression with a spatial field, random community intercept and a random household intercept.

|  | Univariate | Multivariable |
| --- | --- | --- |
| **Individual level** | Odds ratio<br>(95% confidence interval) | Adjusted odds ratio<br>(95% confidence interval) |
| Age group (years): |  |  |
| <5 | 0.00 (0.00 - 4.11e7) | 0.00 (0.00 - 6.42e6) |
| 5-10 | 1.06 (0.39 - 3.11) | 1.10 (0.39 - 3.10) |
| 11-20 | REF | REF |
| 21-30 | 1.04 (0.45 - 2.56) | 0.80 (0.33 - 1.94) |
| 31-40 | 0.99 (0.38 - 2.74) | 0.82 (0.29 - 2.21) |
| 41-50 | 1.42 (0.57 - 3.78) | 1.35 (0.53 - 3.46) |
| 51-60 | 2.05 (0.83 - 5.36) | 2.09 (0.76 - 6.88) |
| >60 | 0.98 (0.32 - 3.23) | 0.90 (0.29 - 2.83) |
| Sex: |  |  |
| Female | REF | REF |
| Male | 0.74 (0.42 - 1.49) | 0.55 (0.25 - 1.03) |
| Dengue Status: |  |  |
| Seronegative | REF | REF |
| Seropositive | 1.75 (0.89 - 3.24) | 2.08 (1.01 - 6.52) |
| Last time left community: |  |  |
| ≤180 days | REF | REF |
| >180 days | 1.18 (0.56 - 2.71) | 0.75 (0.39 - 1.42) |
| **Household level** |  |  |
| Household income (Taka): |  |  |
| <10,000 | REF | REF |
| 10,000 - 20,000 | 0.73 (0.37 - 1.46) | 0.82 (0.34 - 1.87) |
| >20,000 | 1.29 (0.59 - 2.81) | 1.79 (0.68 - 4.82) |
| Household head education: |  |  |
| No education | REF | REF |
| Primary school | 0.96 (0.41 - 2.27) | 1.07 (0.41 - 2.84) |
| High school | 0.76 (0.37 - 1.56) | 0.83 (0.36 - 1.90) |
| Higher | 0.63 (0.26 - 1.47) | 0.70 (0.25 - 1.91) |
| Electricity in home: |  |  |
| No | REF | REF |
| Yes | 1.28 (0.33 - 4.98) | 1.31 (0.33 - 5.38) |
| Own home: |  |  |
| No | REF | REF |
| Yes | 0.27 (0.06 – 0.96) | 0.23 (0.04 - 1.00) |
| Own land away from home: |  |  |
| No | REF | REF |
| Yes | 1.20 (0.63 – 2.30) | 0.93 (0.42 - 2.18) |
| Mosquito control: |  |  |
| No | REF | REF |
| Yes | 0.60 (0.22 – 1.51) | 0.59 (0.22 - 1.52) |
| **Community level** |  |  |
| *A. aegypti* captured: |  |  |
| No | REF | REF |
| Yes | 4.14 (0.46 – 39.24) | 5.29 (0.26 - 215.30) |
| *A. albopictus* captured: |  |  |
| No | REF | REF |
| Yes | 1.76 (0.31 – 11.15) | 2.27 (0.16 - 30.23) |
| Population density (log scale): | 0.62 (0.00 – 2.41) | 1.05 (0.39 - 2.68) |

## Notes

### Competing Interest Statement

The authors have declared no competing interest.

### Funding Statement

This study was funded by from ERC 804744 and the CDC. The authors declare no conflicts of interest.

